# A real-world evaluation of the implementation of NLP technology in abstract screening of a systematic review

**DOI:** 10.1101/2022.02.24.22268947

**Authors:** Sara Perlman-Arrow, Noel Loo, Niklas Bobrovitz, Tingting Yan, Rahul K. Arora

## Abstract

The laborious and time-consuming nature of systematic review production hinders the dissemination of up-to-date evidence synthesis. Well-performing natural language processing (NLP) tools for systematic reviews have been developed, showing promise to improve efficiency. However, the feasibility and value of these technologies have not been comprehensively demonstrated in a real-world review. We developed an NLP-assisted abstract screening tool that provides text inclusion recommendations, keyword highlights, and visual context cues. We evaluated this tool in a living systematic review on SARS-CoV-2 seroprevalence, conducting a quality improvement assessment of screening with and without the tool. We evaluated changes to abstract screening speed, screening accuracy, characteristics of included texts, and user satisfaction. The tool improved efficiency, reducing screening time per abstract by 45.9% and decreasing inter-reviewer conflict rates. The tool conserved precision of article inclusion (positive predictive value; 0.92 with tool vs 0.88 without) and recall (sensitivity; 0.90 vs 0.81). The summary statistics of included studies were similar with and without the tool. Users were satisfied with the tool (mean satisfaction score of 4.2*/*5). We evaluated an abstract screening process where one human reviewer was replaced with the tool’s votes, finding that this maintained recall (0.92 one-person, one-tool vs 0.90 two tool-assisted humans) and precision (0.91 vs 0.92) while reducing screening time by 70%. Implementing an NLP tool in this living systematic review improved efficiency, maintained accuracy, and was well-received by researchers, demonstrating the real-world effectiveness of NLP in expediting evidence synthesis.

## 1 Background

Evidence synthesis is crucial to evidence-based decision-making in modern medicine and public health. [1, 2] It is especially important during health emergencies, when the evidence base can rapidly change. The volume of COVID-19 literature exemplifies this, with over 500 000 articles on the matter published as of October 2021. [3] Unfortunately, producing systematic reviews is time-consuming and laborious. The mean time from registration to publication is 67.3 weeks for PROSPERO-registered reviews. [4, 5] Living systematic reviews are designed to circumvent delays and provide up-to-date results, [6, 7] but it is similarly laborious to update these at an adequate frequency. [8] Hence, there is increasing urgency to expedite evidence synthesis methods without sacrificing quality.

Software tools using natural language processing (NLP) systems have been developed to accelerate systematic review methods, [9] many of which target abstract screening. [10, 11, 12, 13, 14, 15] Under 3% of texts screened are typically eligible for inclusion, [4] making it time-consuming to parse through search results, and particularly useful to expedite this stage of the process. NLP-based screening tools classify abstract inclusion or exclusion, and are trained on abstracts labeled by human reviewers. Examples include Rayyan, [14] DistillerSR, [12] and ResearchScreener, [10] which use naive-Bayes or ngram based support-vector machine approaches. Recently, transformer-based NLP models have shown particular promise for text screening. [16] Transformer models are typically pre-trained on large bodies of medical literature, then fine-tuned on a specific screening task. This results in broadly improved performance. [17, 18]

Despite many existing technologies, there is little data on the real-world utility of such screening tools. Most previous reports focus primarily on performance metrics, such as tool precision and recall on abstracts previously unseen by the model. [19, 20] Some studies have assessed efficiency measures, such as impact to reviewer workload or time saved while screening, but these are typically conducted retrospectively, with data from completed reviews. [15, 11, 12, 21] Only one study to date has evaluated these tools in the context of an ongoing review with user interactions. This evaluation involved only one reviewer, was done after traditional screening was completed, and focused exclusively on screening time. [10]

Furthermore, few reports have evaluated the impact of implementing NLP tools into living literature reviews, [13] and none have assessed user-tool interactions or user satisfaction in this context. Living reviews could benefit from a continuous level of screening efficiency and lend themselves well to integration of NLP tools: an initial manual review can yield a large number of screened texts, which could serve as the training set to develop an algorithm to in turn expedite continuous review updates.

SeroTracker conducts a living systematic review of global SARS-CoV-2 seroprevalence and publishes results onto an interactive dashboard (Serotracker.com). [22, 23] Each week, our team screens 800-1000 new abstracts and extracts approximately 30 articles. To optimize the efficiency of our screening process, we developed an NLP-assisted software tool and conducted a quality improvement (QI) project assessing the efficiency changes and usability of integrating this tool into our usual methods. We evaluated changes in the time taken to conduct screening, the accuracy of the screening process, the characteristics of included texts in our overall review, user interactions with the tool, and user satisfaction with the process. Moreover, we assessed different combinations of reviewer and tool pairing to determine how to best improve our screening process. As an evaluation of NLP-based tools in an ongoing living systematic review, our report provides novel and comprehensive evidence regarding the feasibility of NLP for screening and its real-world performance benefits.

## 2 Methods

### 2.1 The SeroTracker systematic review of SARS-CoV-2 seroprevalence

SeroTracker conducts a living systematic review of SARS-CoV-2 seroprevalence that is registered with PROSPERO (CRD42020183634 - version 7 May 2020). We run weekly searches in four literature databases (MEDLINE, EMBASE, Web of Science and Europe PMC) to find relevant peer-reviewed and pre-print literature. Full details on this review are available in previous publications. [23]

Every week, the searches yield approximately 800-1000 new texts, which are uploaded into the Covidence platform for screening. [24] Abstracts are reviewed in duplicate, with first and second reviewers blinded to each others’ votes. All texts whose abstracts receive “include” votes by two reviewers undergo a full-text screen. All thirteen research team members conduct screening and can provide the first vote, and one of six designated second reviewers provides the second vote. This abstract screening process yields 25-40 articles for full-text screening and 20-35 included articles each week.

### 2.2 Development and implementation of an NLP tool into SeroTracker’s processes

This study was conducted as a quality improvement project, following the Plan-Do-Study-Act (PDSA) model, to facilitate and expedite SeroTracker’s screening process while maintaining accuracy. The Conjoint Health Research Ethics Board (CHREB) at the University of Calgary granted us an exemption from the research ethics board review for this QI project. This study followed the Standards for QUality Improvement Reporting Excellence (SQUIRE-2.0).

#### 2.2.1 Plan

In line with process improvement measures at SeroTracker, reviewers were interviewed to assess satisfaction with the screening process. Team members noted its time-consuming nature and identified the following key challenges: difficulty in tracking the number of texts screened, the inability to reverse a vote if they misclicked, and the inability to identify key information at a glance to determine whether a text should be included.

We developed an NLP-enabled tool that adds additional features to Covidence to allow more efficient identification of text eligibility. This tool included (1) an inclusion recommendation indicator, which displays a confidence rating ranging from “not recommended” to “strongly recommended” in the form of a coloured circle beside the abstract title. This was developed using the transformer-based pre-trained NLP model PubMedBERT. [18] We fine-tuned the model on a set of 25,000 previously screened abstracts from the living systematic review. We also included (2) a feature that highlights the Population, Intervention, and Outcome (PIO) abstract components in different colours, using the same model but trained on the EBM-NLP dataset. [25] More details about the tool’s development can be found in Appendix B.

The tool also incorporated features to streamline screening and ameliorate user experience: (3) a screening progress tracker (4) a button to undo a user’s most recent votes on a text (5) a feature that displays abstracts in a way that separates them by section headings (e.g., “Background”, “Methods”, etc.) and (6) a feature highlighting reviewer-specified keywords (Appendix Figure C3).

#### 2.2.2 Do

We conducted a project with AB design to assess the feasibility and impact of tool implementation on abstract screening. We selected a set of 400 abstracts (“pilot abstracts”) to evaluate tool performance. These abstracts had been previously screened using the same inclusion criteria as part of SeroTracker’s review. 309/400 were previously excluded and 91/400 were previously included in the review.

This project was conducted over a five-week period in three stages Figure 1. In the first two weeks, team members conducted screening without the tool (“without-tool stage”). 200 pilot abstracts were added to the regular primary searches each week. We subsequently implemented a week-long washout period, where no pilot abstracts were added, and reviewers installed and familiarized themselves with the tool. In the final two weeks, team members used the tool for screening (“with-tool stage”) using the features they felt were most helpful. In this stage, 200 pilot abstracts were again added to the regular primary searches each week.

**Figure 1:**
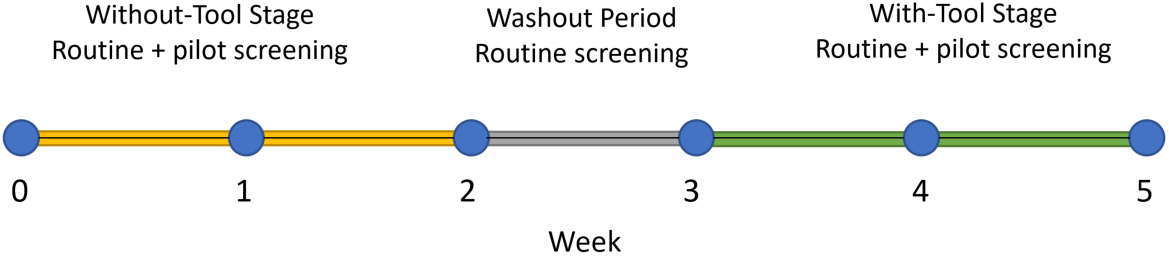
The timeline of the quality improvement project

#### 2.2.3 Study

Three sets of reviewer votes on the pilot abstracts were collected: votes from the initial screen in April (“pre-project votes”), votes from the without-tool, and votes from the with-tool stages. We used pre-project votes as the reference standard for comparison.

We evaluated key process, outcome, and structure measures. Process measures included (1) efficiency metrics, including the screening time and the conflict rate with and without the tool and (2) accuracy metrics, including precision (positive predictive value) and recall (sensitivity) of screening, as well as the performance of the tool’s inclusion recommendations. To evaluate precision and recall, we first calculated the baseline expected variability due to human error in screening, by comparing the included and excluded texts between the pre-project and without-tool stages, as there is inherent human error in the systematic review process. [26] We then assessed whether the outcomes of the with-tool stage were within expected levels of variability.

The first outcome measure evaluated was the tool’s impact on results of the review, assessed by comparing summary descriptive statistics for included seroprevalence estimates in the pre-project, without-tool, and with-tool stages. We also assessed reviewers’ usage of different features, such as voter alignment with the NLP recommendations and the frequency of use of each feature. We surveyed users to understand overall satisfaction with the tool.

Finally, one structure measure was evaluated, which compared tool performance with different combinations of human and tool votes. We assessed the tool’s performance using data from the project in a “one-person and one-tool” (OPOT) screening process, a simulated abstract screening scenario in which one human reviewer is replaced with the tool’s automated inclusion recommendations. The tool voted “include” on an abstract if it provided a rating of “weakly recommended” or stronger for the abstract. Two scenarios were considered: one in which the human reviewer had access to the tool [OPOT (W)], and one in which they did not [OPOT (W/O)]. Each of these scenarios was compared to a “toolonly” system, in which only the tool’s automated inclusion recommendations were used for abstract screening, maintaining conflict resolution and full-text screen processes between human reviewers. For the human reviewer screening scenarios, we used human votes from the without and with-tool stages from the voter with the longest tenure at SeroTracker. Any additional conflict resolution or full-text screenings required for this analysis were done after the with-tool stage by human reviewers.

Details about the process, outcome, and structure measures studied are presented in Table 1, along with their key results.

**Table 1:**
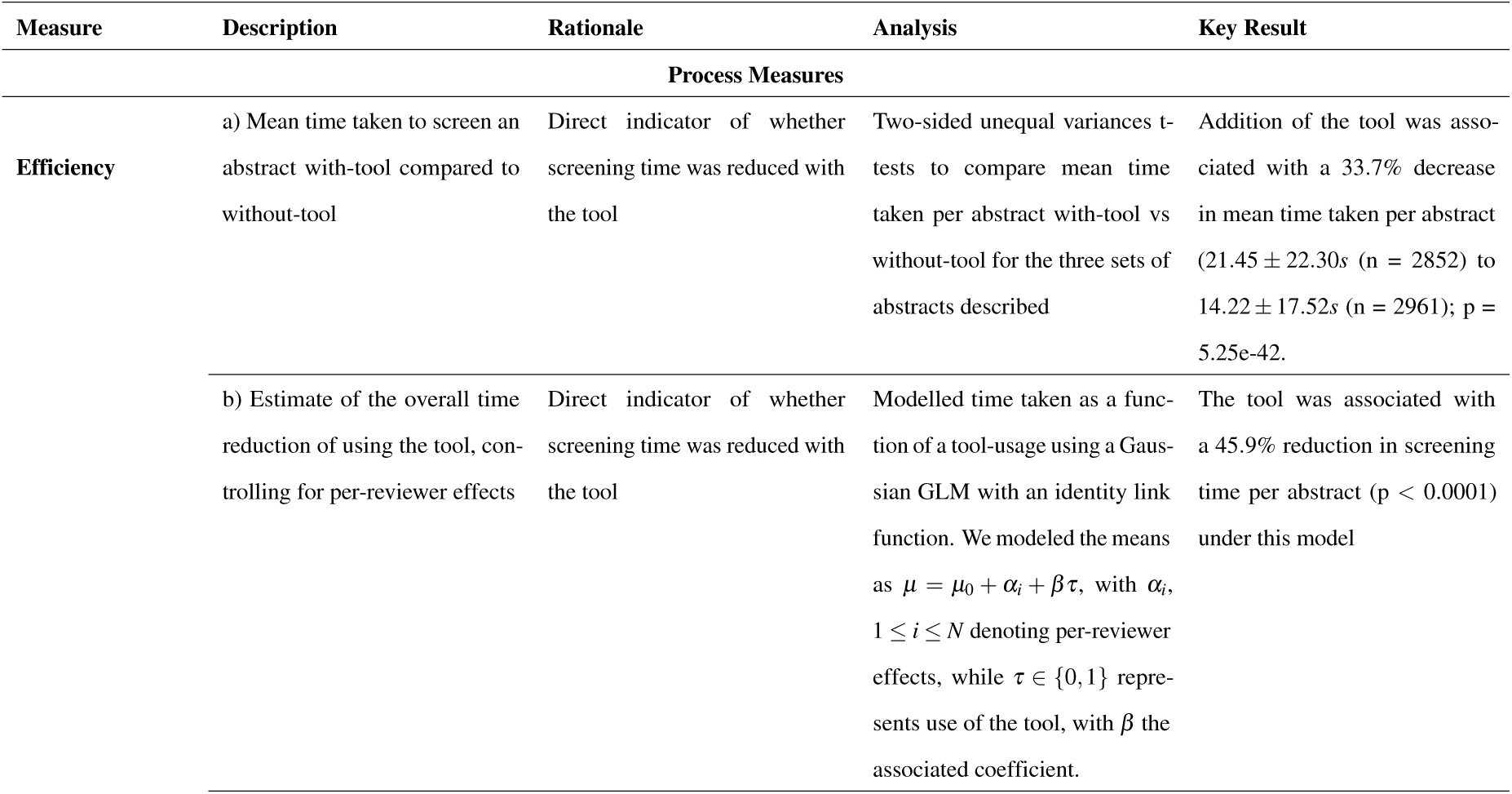
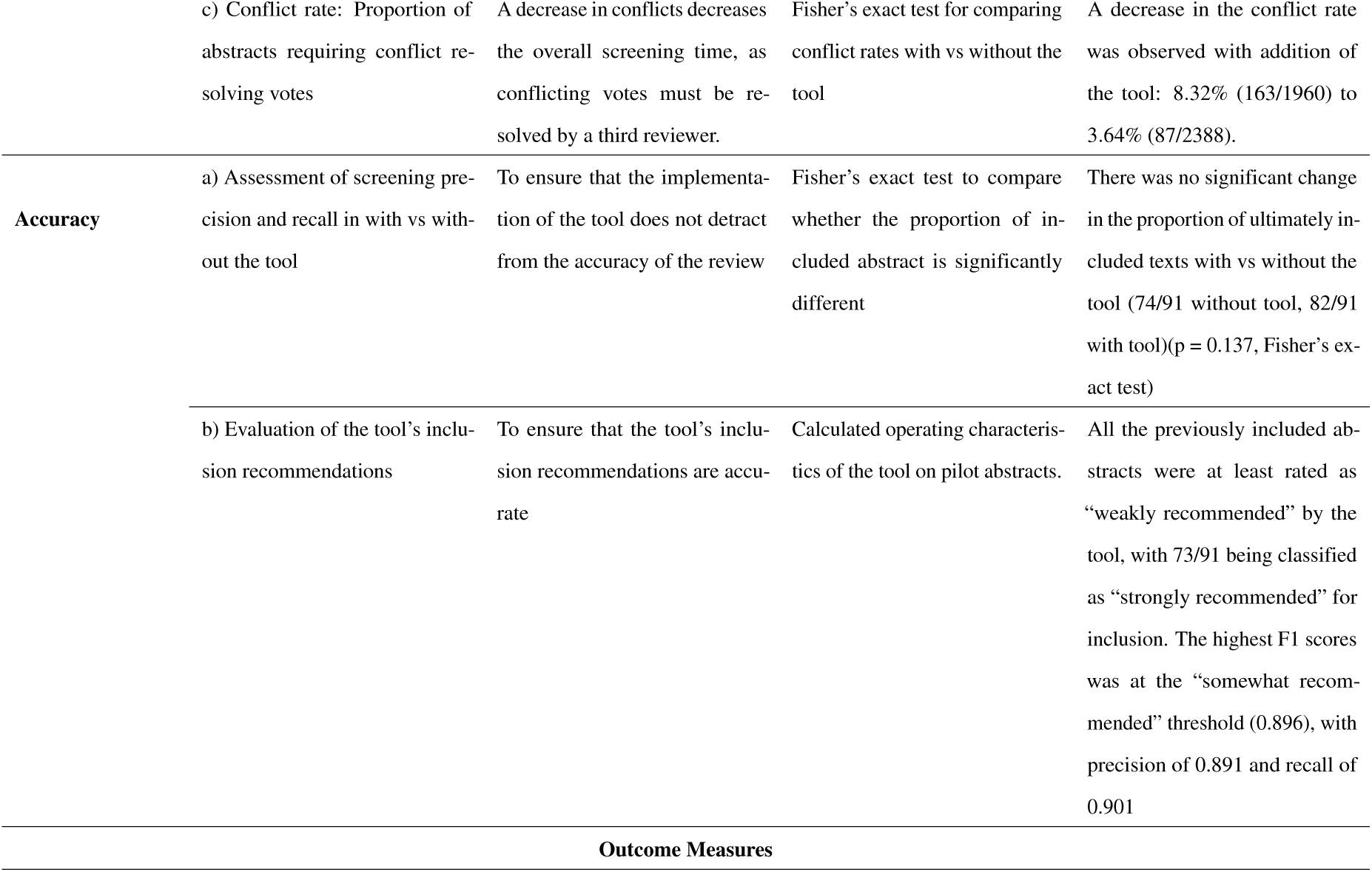
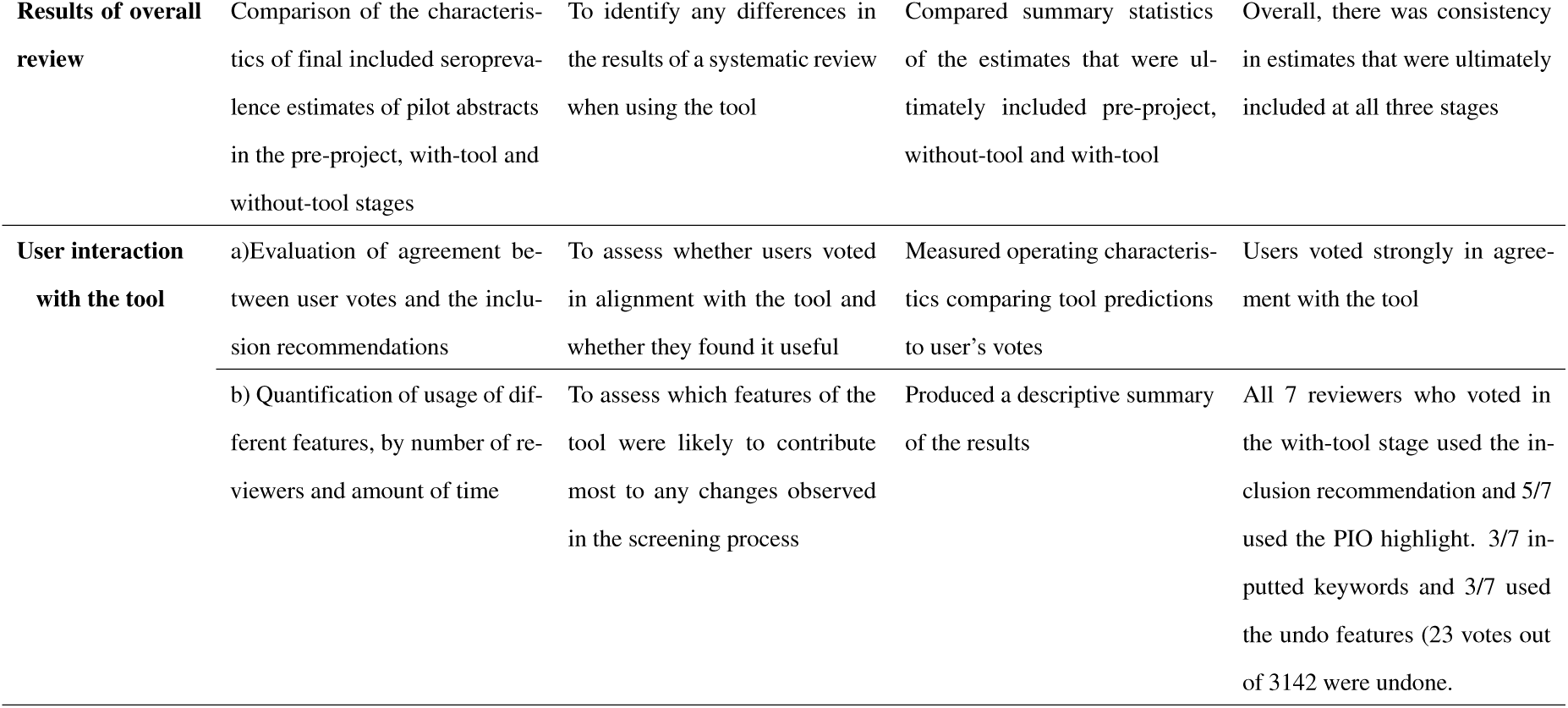
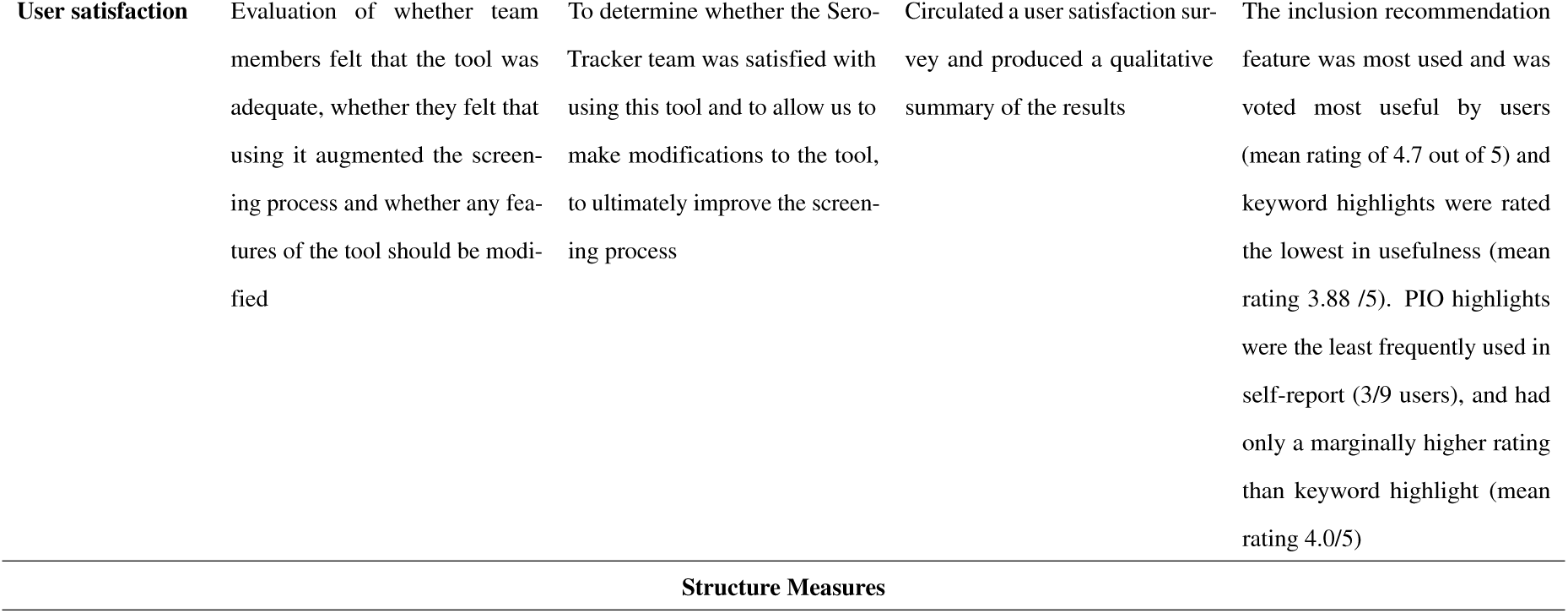
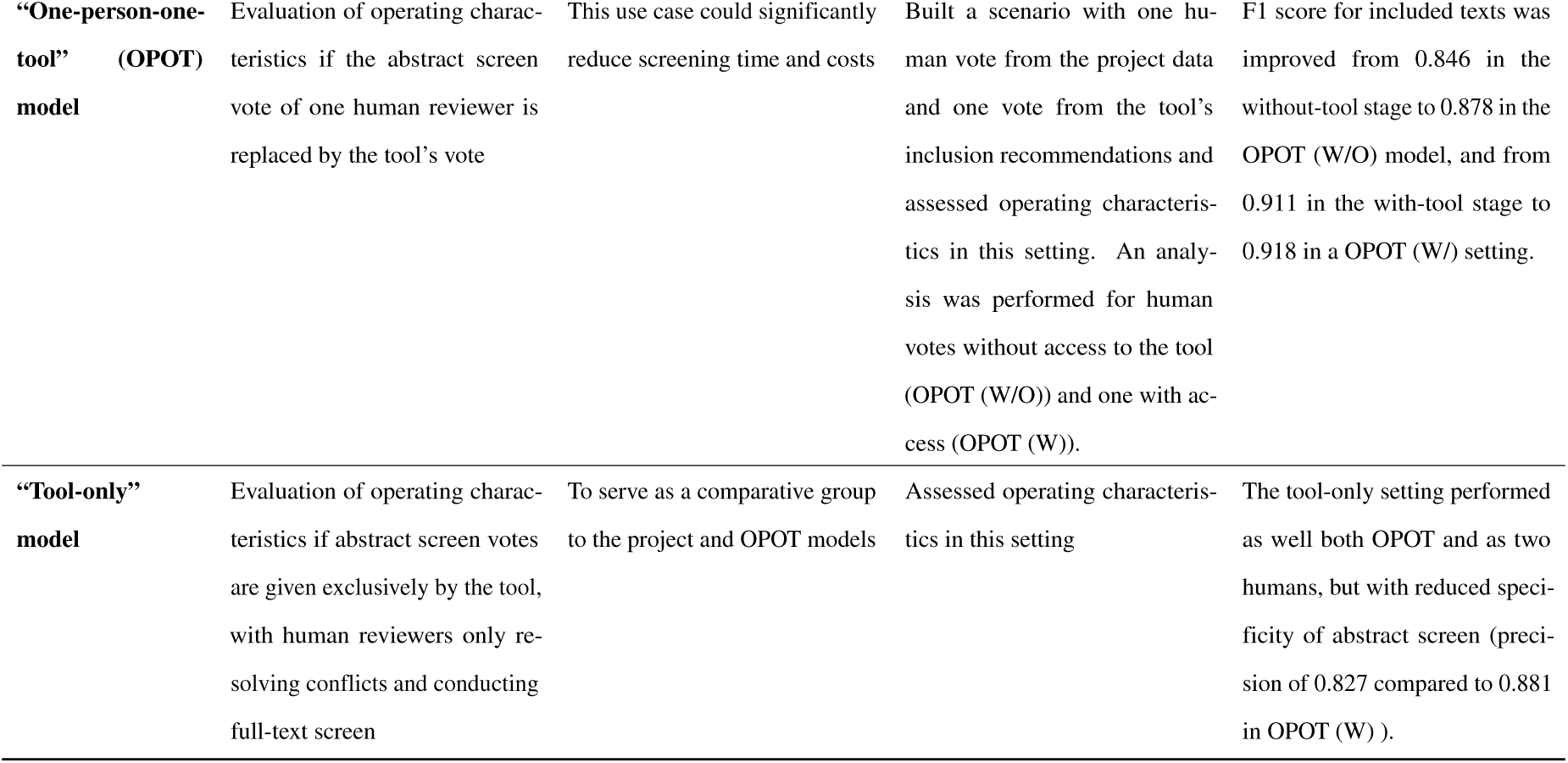
Key methods and results of process, outcome and structure measures evaluated.

| Measure | Description | Rationale | Analysis | Key Result |
| --- | --- | --- | --- | --- |
| Process Measures |  |  |  |  |
| Efficiency | a) Mean time taken to screen an abstract with-tool compared to without-tool | Direct indicator of whether screening time was reduced with the tool | Two-sided unequal variances t-tests to compare mean time taken per abstract with-tool vs without-tool for the three sets of abstracts described | Addition of the tool was associated with a 33.7% decrease in mean time taken per abstract ($21.45 \pm 22.30s$ (n = 2852) to $14.22 \pm 17.52s$ (n = 2961); p = 5.25e-42. |
| | b) Estimate of the overall time reduction of using the tool, controlling for per-reviewer effects | Direct indicator of whether screening time was reduced with the tool | Modelled time taken as a function of a tool-usage using a Gaussian GLM with an identity link function. We modeled the means as $\mu = \mu_0 + \alpha_i + \beta\tau$, with $\alpha_i$, $1 \leq i \leq N$ denoting per-reviewer effects, while $\tau \in \{0,1\}$ represents use of the tool, with $\beta$ the associated coefficient. | The tool was associated with a 45.9% reduction in screening time per abstract (p < 0.0001) under this model |
|  | c) Conflict rate: Proportion of abstracts requiring conflict resolving votes | A decrease in conflicts decreases the overall screening time, as conflicting votes must be resolved by a third reviewer. | Fisher's exact test for comparing conflict rates with vs without the tool | A decrease in the conflict rate was observed with addition of the tool: 8.32% (163/1960) to 3.64% (87/2388). |
| **Accuracy** | a) Assessment of screening precision and recall in with vs without the tool | To ensure that the implementation of the tool does not detract from the accuracy of the review | Fisher's exact test to compare whether the proportion of included abstract is significantly different | There was no significant change in the proportion of ultimately included texts with vs without the tool (74/91 without tool, 82/91 with tool)(p = 0.137, Fisher's exact test) |
|  | b) Evaluation of the tool's inclusion recommendations | To ensure that the tool's inclusion recommendations are accurate | Calculated operating characteristics of the tool on pilot abstracts. | All the previously included abstracts were at least rated as "weakly recommended" by the tool, with 73/91 being classified as "strongly recommended" for inclusion. The highest F1 scores was at the "somewhat recommended" threshold (0.896), with precision of 0.891 and recall of 0.901 |
| **Outcome Measures** |  |  |  |  |
| **Results of overall review** | Comparison of the characteristics of final included seroprevalence estimates of pilot abstracts in the pre-project, with-tool and without-tool stages | To identify any differences in the results of a systematic review when using the tool | Compared summary statistics of the estimates that were ultimately included pre-project, without-tool and with-tool | Overall, there was consistency in estimates that were ultimately included at all three stages |
| **User interaction with the tool** | a) Evaluation of agreement between user votes and the inclusion recommendations | To assess whether users voted in alignment with the tool and whether they found it useful | Measured operating characteristics comparing tool predictions to user's votes | Users voted strongly in agreement with the tool |
|  | b) Quantification of usage of different features, by number of reviewers and amount of time | To assess which features of the tool were likely to contribute most to any changes observed in the screening process | Produced a descriptive summary of the results | All 7 reviewers who voted in the with-tool stage used the inclusion recommendation and 5/7 used the PIO highlight. 3/7 inputted keywords and 3/7 used the undo features (23 votes out of 3142 were undone). |
| **User satisfaction** | Evaluation of whether team members felt that the tool was adequate, whether they felt that using it augmented the screening process and whether any features of the tool should be modified | To determine whether the SeroTracker team was satisfied with using this tool and to allow us to make modifications to the tool, to ultimately improve the screening process | Circulated a user satisfaction survey and produced a qualitative summary of the results | The inclusion recommendation feature was most used and was voted most useful by users (mean rating of 4.7 out of 5) and keyword highlights were rated the lowest in usefulness (mean rating 3.88 /5). PIO highlights were the least frequently used in self-report (3/9 users), and had only a marginally higher rating than keyword highlight (mean rating 4.0/5) |
| Structure Measures |  |  |  |  |
| **“One-person-one-tool” (OPOT) model** | Evaluation of operating characteristics if the abstract screen vote of one human reviewer is replaced by the tool’s vote | This use case could significantly reduce screening time and costs | Built a scenario with one human vote from the project data and one vote from the tool’s inclusion recommendations and assessed operating characteristics in this setting. An analysis was performed for human votes without access to the tool (OPOT (W/O)) and one with access (OPOT (W)). | F1 score for included texts was improved from 0.846 in the without-tool stage to 0.878 in the OPOT (W/O) model, and from 0.911 in the with-tool stage to 0.918 in a OPOT (W/) setting. |
| **“Tool-only” model** | Evaluation of operating characteristics if abstract screen votes are given exclusively by the tool, with human reviewers only resolving conflicts and conducting full-text screen | To serve as a comparative group to the project and OPOT models | Assessed operating characteristics in this setting | The tool-only setting performed as well both OPOT and as two humans, but with reduced specificity of abstract screen (precision of 0.827 compared to 0.881 in OPOT (W) ). |

#### 2.2.4 Act

Results from this work were used to inform whether to integrate this tool into regular practice at SeroTracker and to inform further improvements.

## 3 Results

### 3.1 Process measures

#### 3.1.1 Screening efficiency - time taken to screen

Across all abstracts, the tool was associated with a 33.7% decrease in mean screening time per abstract from 21.45 *±* 22.30s (n = 2852) to 14.22 *±* 17.52s (n = 2961) (p = 5.25e-42 by two-sided unequal-variances t-test) (Table 2, Figure 2a).

**Table 2:**
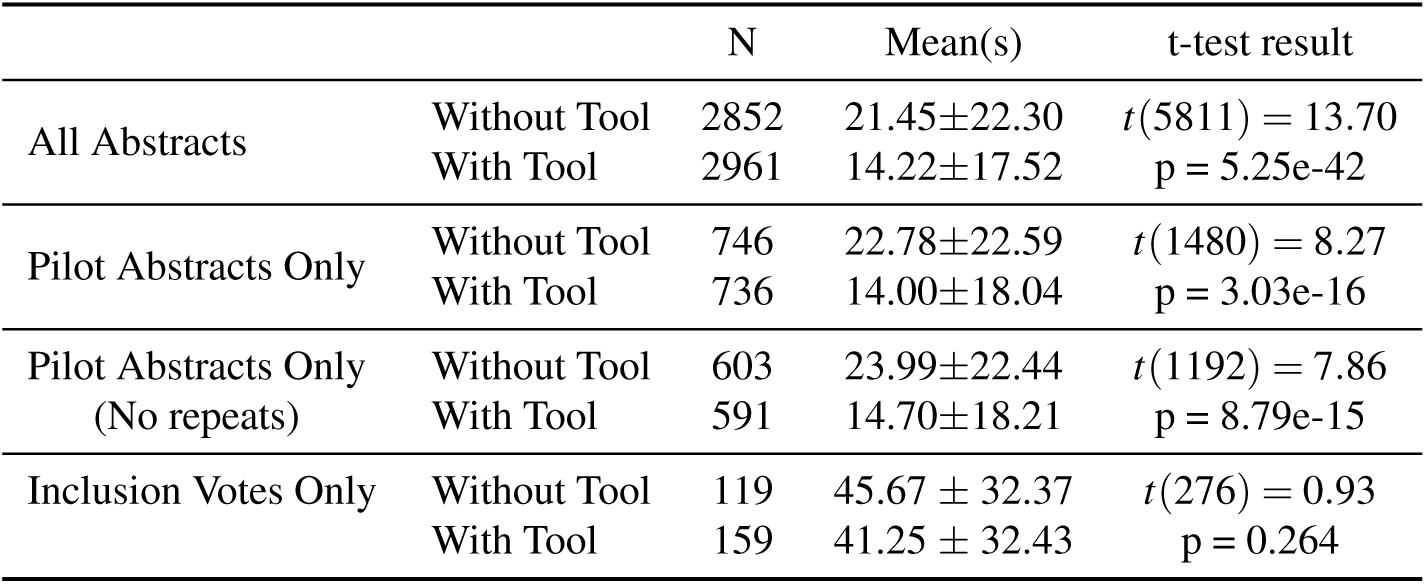
Mean screening time per abstract with-tool and without-tool, stratified into four categories: all abstracts, pilot abstracts only, pilot abstracts excluding repeated reviewer-abstract vote pairs, and only abstracts with “include” vote.

**Figure 2:**
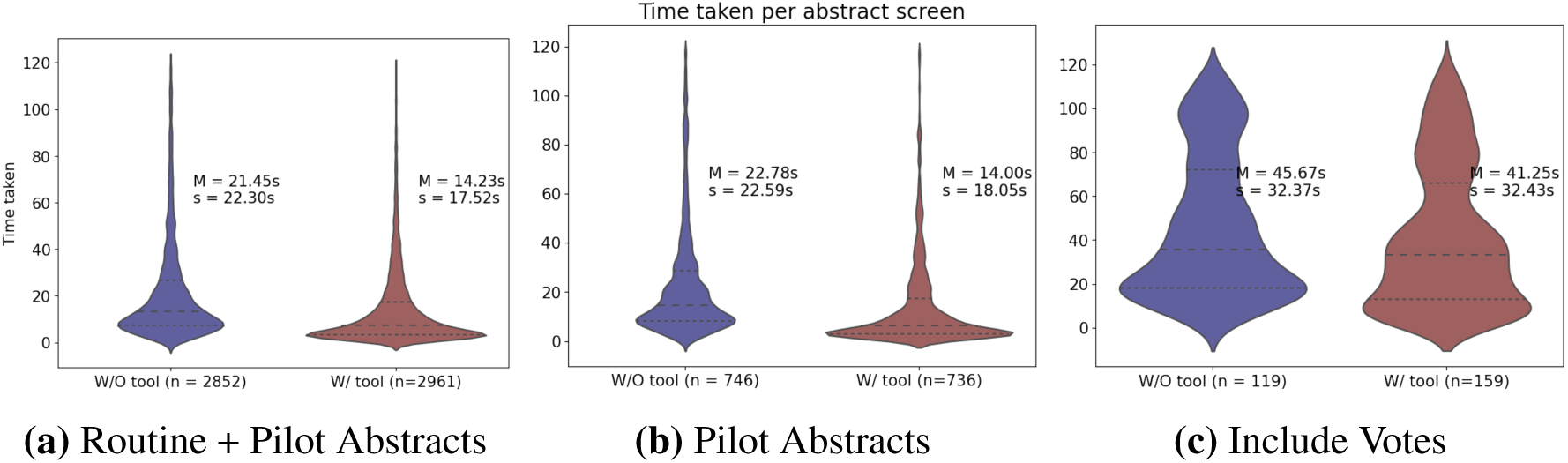
Violin plots of times taken per abstract screen with and without the tool. Results are split into three categories: (a) all abstracts, (b) pilot abstracts only, and (c) abstracts which received an inclusion vote. Means and standard deviations are reported by the black and grey dotted lines, respectively.

This reduction was similar when considering the pilot abstracts alone (22.78 *±* 22.59s (n = 746) to 14.00 *±* 18.04s (n = 736); p = 3.03e-16) (Table 2, Figure 2b).168/800 pilot abstracts were screened twice by the same reviewer with and without the tool. To account for possible order effects in these votes, we repeated this analysis excluding them. We continued to observe a significant decrease in abstract screening time, from 23.99 *±* 22.44s without-tool (n = 603) to 14.70 *±* 18.21s with-tool (n = 591) (p = 8.79e-15) (Table 2).

Lastly, we repeated the analysis only using abstracts ultimately included at the abstract screening stage. These took a similar time to screen with or without the tool (Table 2, Figure 2c).

To account for inter-reviewer effects and inter-abstract variability in our analysis of the tool, we modeled the distribution of the logarithm of the times taken as a function of tool-use, adjusted for effects of reviewer speed. Under this model, the tool was associated with a 45.9% reduction in screening time per abstract (p < 0.0001). There was substantial variability in the mean screening time for different reviewers, with *a*_*i*_ ranging from -0.96 to 0.66.

The tool’s impact on conflict rate was also assessed. An increased number of conflicting votes decreases the efficiency of abstract screening, as a third reviewer must resolve these. Of the 1960 unique abstracts voted on without the tool, 163 had a conflicting vote. The conflict rate decreased from 8.32% (163/1960) to 3.64% (87/2388) (p=5.36e-11, Fisher’s exact test) when the tool was added (Table 3). When considering only the pilot abstracts, a similar decrease was observed: 11.25% (45/400) to 5.75% (23/400) (p = 0.007, Fisher’s exact test) (Table 3). This conflict rate reduction results in a 2.2% time savings per abstract.

**Table 3:** Conflict rate of abstract screening with-tool vs without-tool, for all abstracts and for pilot abstracts only.

|  |  | Conflicts | Abstracts | Rate | Test result |
| --- | --- | --- | --- | --- | --- |
| All Abstracts | Without Tool | 163 | 1960 | 8.32% | $p = 5.36e-11$ |
|  | With Tool | 2961 | 2388 | 3.64% |  |
| Pilot Abstracts | Without Tool | 45 | 400 | 11.25% | $p = 0.007$ |
|  | With Tool | 23 | 400 | 5.75% |  |

In summary, the tool improved all efficiency metrics except the mean time taken for included abstracts. Combining the conflict rate savings (2.2%) and mean times savings (33.7%), we expect a 35.2% reduction in screening time per abstract, leading to 3.93 hours of reviewer time saved each week.

#### 3.1.2 Effect on screening precision and recall

We compared the precision and recall of the screening process with and without the tool to ensure that tool use did not interfere with screening accuracy. When considering pilot abstracts that were included past full-text review, 74 of the 91 previously included abstracts (PI) were included without the tool and 82 with it (Table 4). This change was not significant (p = 0.137, Fisher’s exact test), meaning accuracy was conserved. Looking at the texts included past full-text review, there were 7 false positive (FP) texts with the tool and 10 without it. All FPs were found to have been excluded in the extraction stage. This means that no new texts were included in the review as a result of the project. ^1^.

**Table 4:** Precision, recall and F1 scores in all user-tool pairings for texts that were included in abstract screening and full text screening stages

|  |  | TN | FP | FN | TP | Prec. | Rec. | F1 |
| --- | --- | --- | --- | --- | --- | --- | --- | --- |
| Included Abstracts | Without | 297 | 12 | 14 | 77 | 0.865 | 0.846 | 0.856 |
|  | With | 298 | 11 | 3 | 88 | 0.889 | 0.967 | 0.926 |
|  | OPOT(W/O) | 298 | 11 | 7 | 84 | 0.884 | 0.923 | 0.903 |
|  | OPOT(W) | 297 | 12 | 2 | 89 | 0.881 | 0.978 | 0.927 |
|  | Tool-Only | 290 | 19 | 0 | 91 | 0.827 | 1.000 | 0.905 |
| Included Full Texts | Without | 299 | 10 | 17 | 74 | 0.881 | 0.813 | 0.846 |
|  | With | 302 | 7 | 9 | 82 | 0.921 | 0.901 | 0.911 |
|  | OPOT(W/O) | 299 | 10 | 12 | 79 | 0.888 | 0.868 | 0.878 |
|  | OPOT(W) | 301 | 8 | 7 | 84 | 0.913 | 0.923 | 0.918 |
|  | Tool-Only | 301 | 8 | 6 | 85 | 0.914 | 0.934 | 0.924 |

Abbreviations: TN, true negative ; FP, false positive ; FN, false negative ; TP, true positive ; Prec., precision ; Rec., recall

#### 3.1.3 Inclusion recommendation performance on the pilot abstracts

The tool’s inclusion recommendation feature rates each abstract’s inclusion likelihood into one of five categories. The feature’s operating characteristics were evaluated on the pilot abstracts (Table 5, Figure 3). FPs and FNs were calculated taking the pre-project votes as “true”, and an inclusion prediction was assigned if the tool recommended at least that level of confidence. All the PI pilot abstracts were at least weakly recommended, with 73/91 being strongly recommended. The weakly recommended threshold gave the highest F1 score (0.905), with a precision of 0.827 and recall of 1.0 at this level. Scores remained high at all thresholds.

**Table 5:** Operating characteristics of the tool evaluated on the pilot abstracts at different inclusion thresholds.

|  | TN | FP | FN | TP | Prec. | Rec. | F1 |
| --- | --- | --- | --- | --- | --- | --- | --- |
| Not Recommended | 0 | 309 | 0 | 91 | 0.228 | 1.000 | 0.371 |
| Weakly Recommended | 290 | 19 | 0 | 91 | 0.827 | 1.000 | 0.905 |
| Somewhat Recommended | 299 | 10 | 9 | 82 | 0.891 | 0.901 | 0.896 |
| Recommended | 301 | 8 | 14 | 77 | 0.906 | 0.846 | 0.875 |
| Strongly Recommended | 302 | 7 | 18 | 73 | 0.912 | 0.802 | 0.854 |

Abbreviations: TN, true negative ; FP, false positive ; FN, false negative ; TP, true positive ; Prec., precision ; Rec., recall

**Figure 3:**
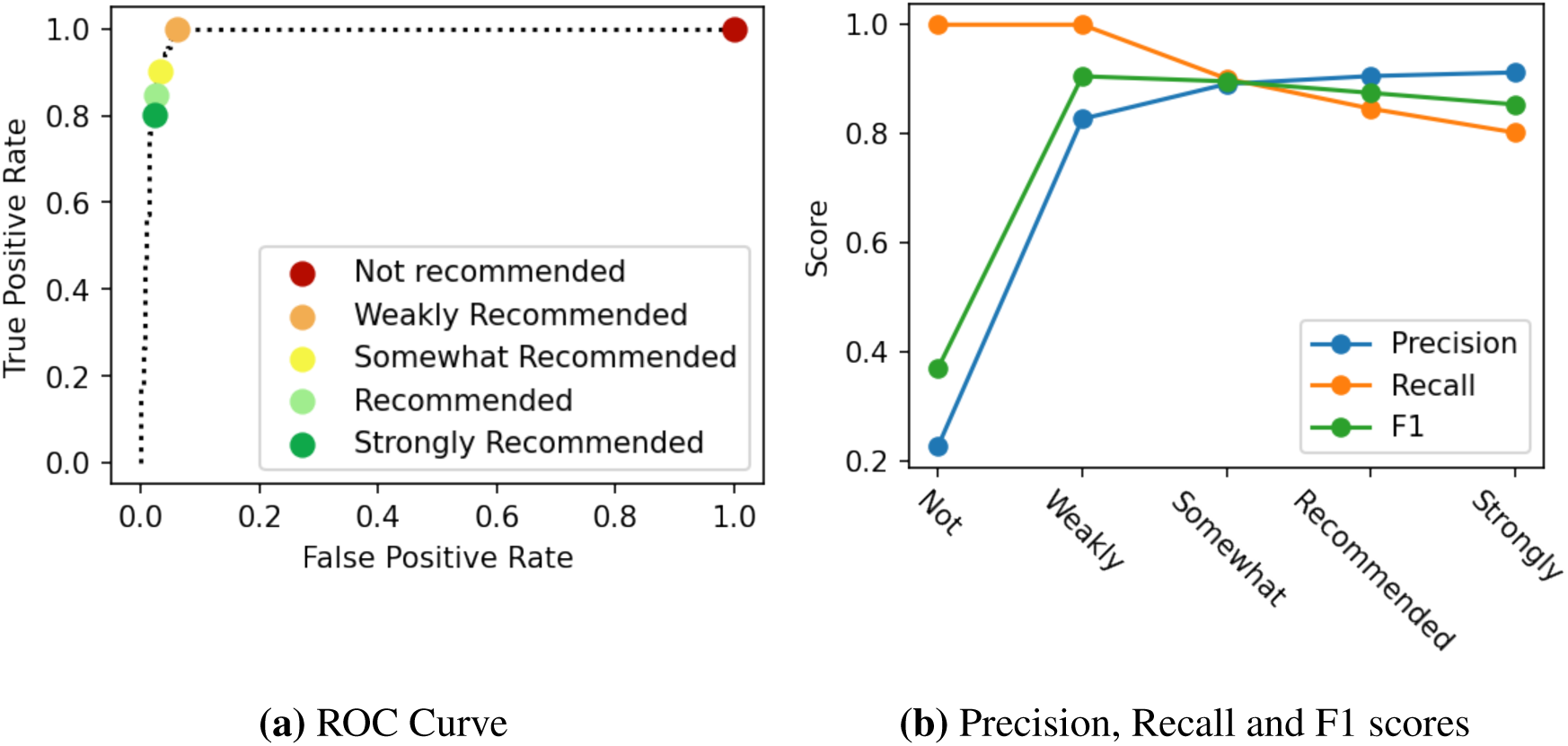
Operating characteristics of the tool evaluated on the pilot abstracts, with “True” labels taken as the outcome of the previous full screening, and the predicted labels taken as the tool’s inclusion likelihood. (a) shows the ROC curve, with the four confidence thresholds given by the tool marked on the curve. (b) shows the precision, recall and F1 scores as a function of the tool’s confidence threshold, from the lower confidence (at least not recommended/red) to the highest (at least strongly recommended/dark green).

### 3.2 Outcome measures

#### 3.2.1 Results of overall systematic review

Table 6 summarizes the statistics of the seroprevalence estimates included in the pre-project stage, the without-tool stage, and the with-tool stage. The majority of the statistics remain consistent with and without the tool. Compared to the pre-project votes, the with-tool stage did not exclude any estimates deemed to have a low or moderate risk of bias.

**Table 6:**
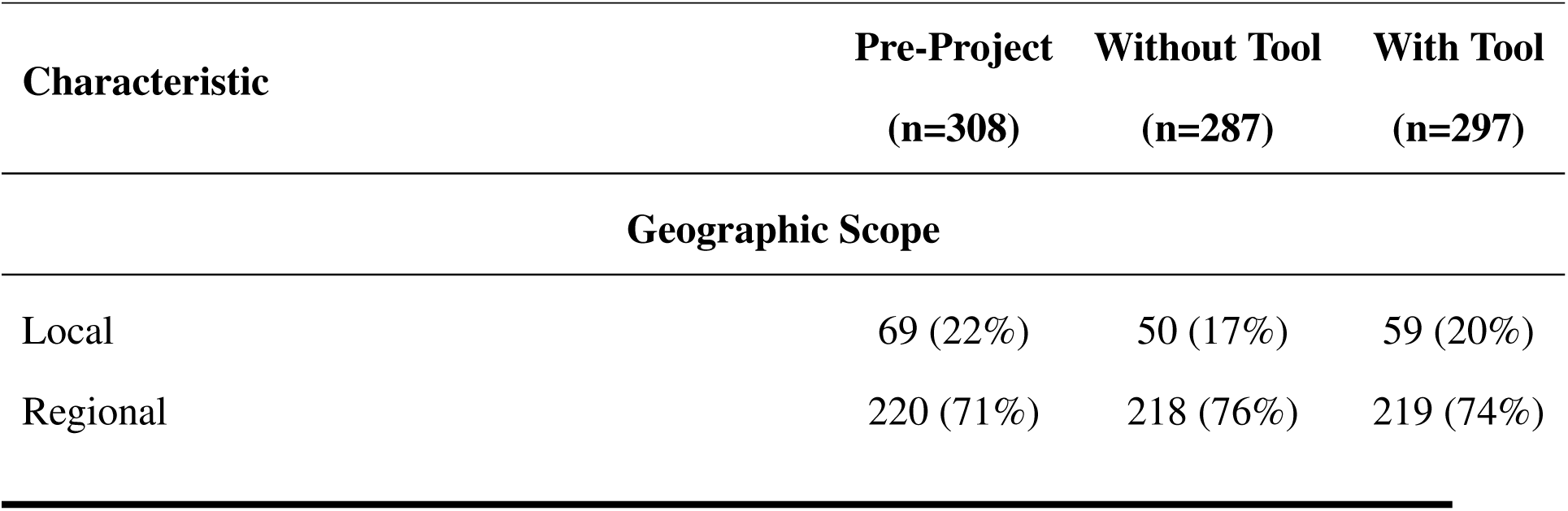
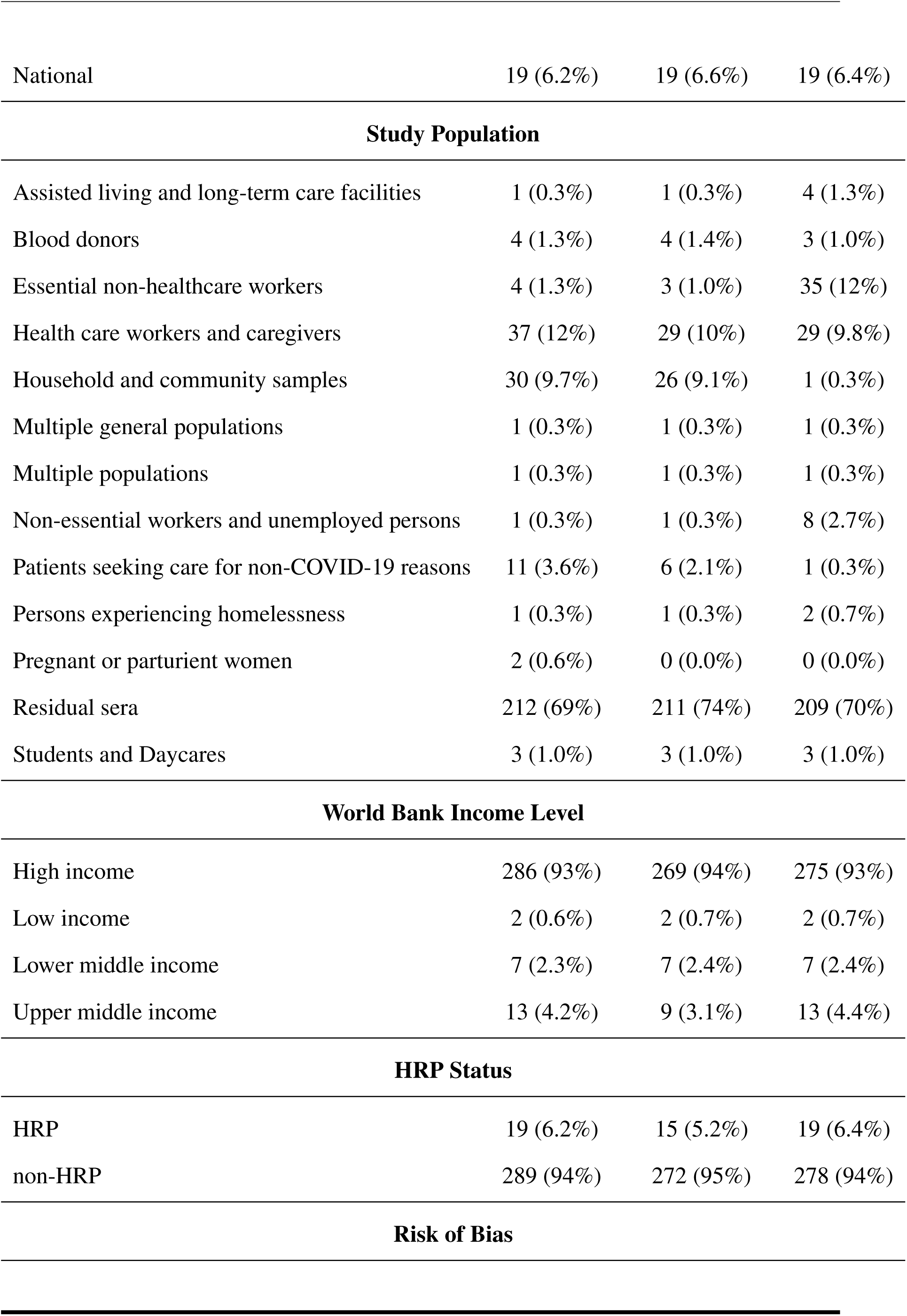
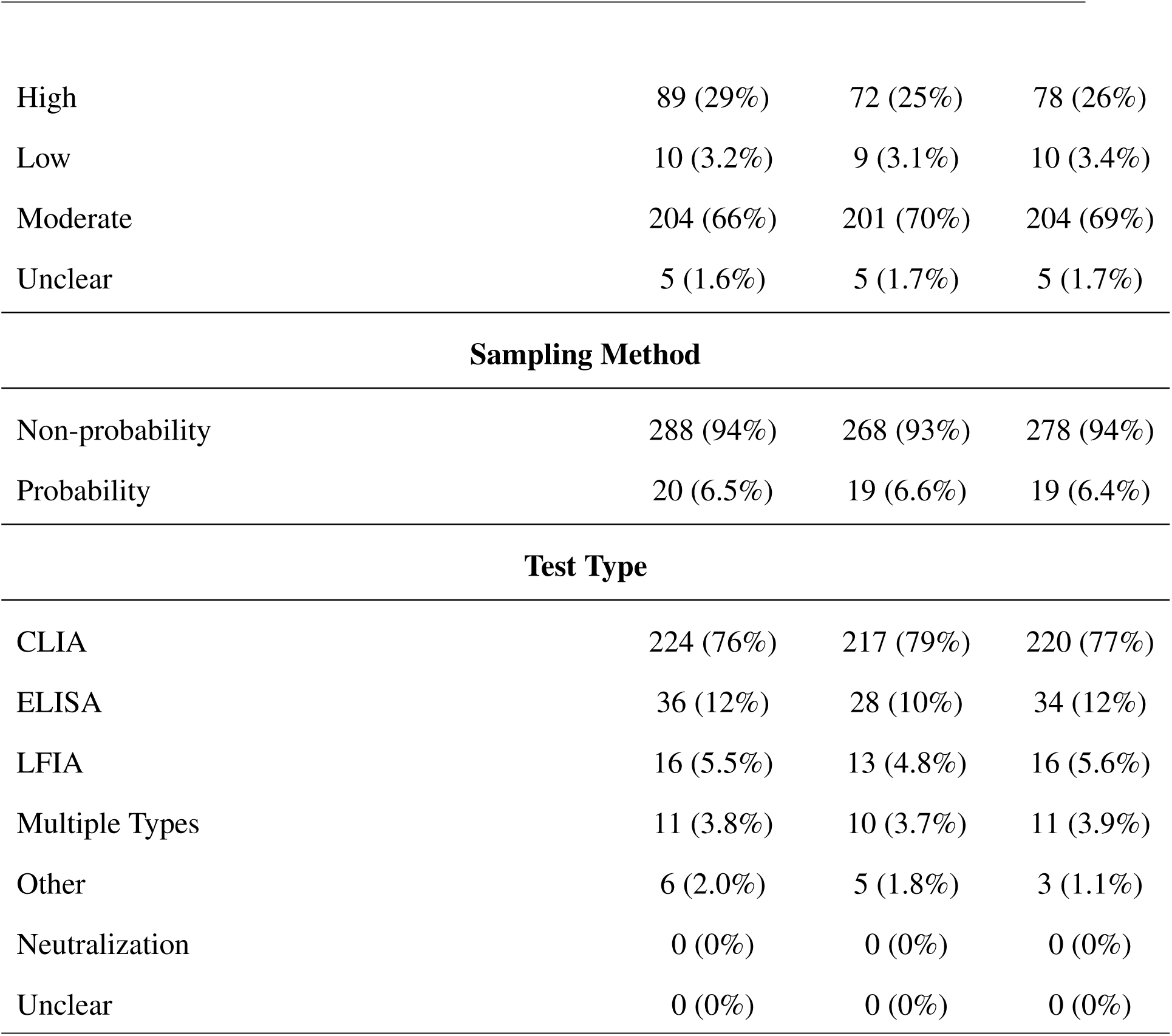
Summary statistics of the seroprevalence estimates from the included texts in the pre-project, without-tool and with-tool stages.

| Characteristic | Pre-Project<br>(n=308) | Without Tool<br>(n=287) | With Tool<br>(n=297) |
| --- | --- | --- | --- |
| **Geographic Scope** |  |  |  |
| Local | 69 (22%) | 50 (17%) | 59 (20%) |
| Regional | 220 (71%) | 218 (76%) | 219 (74%) |
| National | 19 (6.2%) | 19 (6.6%) | 19 (6.4%) |
| **Study Population** |  |  |  |
| Assisted living and long-term care facilities | 1 (0.3%) | 1 (0.3%) | 4 (1.3%) |
| Blood donors | 4 (1.3%) | 4 (1.4%) | 3 (1.0%) |
| Essential non-healthcare workers | 4 (1.3%) | 3 (1.0%) | 35 (12%) |
| Health care workers and caregivers | 37 (12%) | 29 (10%) | 29 (9.8%) |
| Household and community samples | 30 (9.7%) | 26 (9.1%) | 1 (0.3%) |
| Multiple general populations | 1 (0.3%) | 1 (0.3%) | 1 (0.3%) |
| Multiple populations | 1 (0.3%) | 1 (0.3%) | 1 (0.3%) |
| Non-essential workers and unemployed persons | 1 (0.3%) | 1 (0.3%) | 8 (2.7%) |
| Patients seeking care for non-COVID-19 reasons | 11 (3.6%) | 6 (2.1%) | 1 (0.3%) |
| Persons experiencing homelessness | 1 (0.3%) | 1 (0.3%) | 2 (0.7%) |
| Pregnant or parturient women | 2 (0.6%) | 0 (0.0%) | 0 (0.0%) |
| Residual sera | 212 (69%) | 211 (74%) | 209 (70%) |
| Students and Daycares | 3 (1.0%) | 3 (1.0%) | 3 (1.0%) |
| **World Bank Income Level** |  |  |  |
| High income | 286 (93%) | 269 (94%) | 275 (93%) |
| Low income | 2 (0.6%) | 2 (0.7%) | 2 (0.7%) |
| Lower middle income | 7 (2.3%) | 7 (2.4%) | 7 (2.4%) |
| Upper middle income | 13 (4.2%) | 9 (3.1%) | 13 (4.4%) |
| **HRP Status** |  |  |  |
| HRP | 19 (6.2%) | 15 (5.2%) | 19 (6.4%) |
| non-HRP | 289 (94%) | 272 (95%) | 278 (94%) |
| **Risk of Bias** |  |  |  |
| High | 89 (29%) | 72 (25%) | 78 (26%) |
| Low | 10 (3.2%) | 9 (3.1%) | 10 (3.4%) |
| Moderate | 204 (66%) | 201 (70%) | 204 (69%) |
| Unclear | 5 (1.6%) | 5 (1.7%) | 5 (1.7%) |

| Sampling Method |  |  |  |
| --- | --- | --- | --- |
| Non-probability | 288 (94%) | 268 (93%) | 278 (94%) |
| Probability | 20 (6.5%) | 19 (6.6%) | 19 (6.4%) |

| Test Type |  |  |  |
| --- | --- | --- | --- |
| CLIA | 224 (76%) | 217 (79%) | 220 (77%) |
| ELISA | 36 (12%) | 28 (10%) | 34 (12%) |
| LFIA | 16 (5.5%) | 13 (4.8%) | 16 (5.6%) |
| Multiple Types | 11 (3.8%) | 10 (3.7%) | 11 (3.9%) |
| Other | 6 (2.0%) | 5 (1.8%) | 3 (1.1%) |
| Neutralization | 0 (0%) | 0 (0%) | 0 (0%) |
| Unclear | 0 (0%) | 0 (0%) | 0 (0%) |

#### 3.2.2 User interaction with the tool

We first evaluated the agreement between user votes and the tool’s five thresholds of inclusion recommendations. It was found that users typically agreed with the recommendations, with the highest F1 score (0.821) being at the “somewhat recommended” threshold (Appendix A.1) There was a positive correlation between users voting”include” and the presence of keywords in the abstract (Appendix A.2).

The inclusion recommendation feature was the most commonly used. It was used by all 7 unique reviewers in the with-tool stage and in 3140/3142 unique votes. PIO highlighting was used by 5/7 reviewers, and 1918/3142 (61.0%) votes. While all users had the keyword feature on, only 3 actually had any keywords specified, resulting in 21.1% (663/3142) of abstract votes having the features on with keywords input. The undo feature was the least used (3/7 reviewers ; 23/3142 texts), with the vote being changed in only 6 cases. Of note, by default undoing a vote in Covidence is not always possible.

#### 3.2.3 User feedback on the tool

After the project’s conclusion, reviewers were sent a satisfaction survey. 9 of 13 members provided feedback (Table 7). The self-reported usage information did not align perfectly with the computer recorded usage. This could be due to recall bias, as the survey was conducted one week after the conclusion of the project. Reviewers who used features rated their usefulness out of five (Table 7). The inclusion recommendation feature was voted the most useful (mean score of 4.70/5), and keyword (3.88/5) and PIO highlighting (4.00/5) were the least useful.

**Table 7:** Self reported usage and usefulness rating by 9 users for features of the NLP-based tool.

| Feature | Self-reported Usage | Mean usefulness |
| --- | --- | --- |
| Undo | 4/9 | 4.4 (3,4,5,5,5) |
| Inclusion Recommendations | 7/9 | 4.7 (4,4,5,5,5,5,5) |
| Bold Headings | 7/9 | 4.13 (3,3,3,4,5,5,5,5) |
| Keyword Highlighting | 7/9 | 3.88 (3,3,3,4,4,4,2,2) |
| PIO Highlighting | 3/9 | 4 (3,3,4,5,5) |

Reviewers reported that the tool improved perceived screening speed by allowing them to rapidly identify key information that qualifies or disqualifies abstracts for inclusion, specifically through the bolding headings, PIO highlighting, and keyword highlighting. While the undo feature was rarely used, users reported that it provided them with more security, allowing them to correct mistakes that would otherwise be permanent. While many users found the inclusion recommendations useful, they noted that it could give a false sense of security and cause users to blindly trust the tool, rather than carefully read through the abstract. Users also noted that the PIO highlighting feature often highlighted incorrect information, making it distracting at times. This complaint was reflected in the low adoption of the PIO highlighting feature.

### 3.3 Structure measures

#### 3.3.1 One-person-one-tool (OPOT) model

Given the reliability of the tool’s inclusion recommendations, we evaluated outcomes of an abstract screening process where one human reviewer was replaced with the tool’s vote, in a scenario in which the human reviewer did not have access to the tool (OPOT (W/O)) and in one where they did (OPOT (W)).

Results are reported in Table 4 for texts that were included in the abstract screen and for texts that were included in the full-text screen. None of the FPs were ultimately included in the review; all texts were excluded during extraction. Recall for included texts was improved from 0.813 without-tool to 0.868 in the OPOT (W/O) model and from 0.901 with-tool to 0.923 in a OPOT (W/) setting. Assuming that conflict rate remains the same in the OPOT (W) setting as with-tool, OPOT would result in a further 48.6% reduction in screening time, for a potential total time saving of 72.2%. Assuming conflict rate is doubled, the additional OPOT savings would be 45.8% - a total time saving of 70.7%.

The tool-only screening scenario, in which votes are provided by the tool while a human reviewer conducts only conflict resolution and full-text screening, performed comparably well to both OPOT and to two humans. Precision, however, was reduced when using this system (Table 4).

## 4 Discussion

SeroTracker improved its screening processes by implementing an NLP-based tool. Tool use significantly reduced both the mean time taken per abstract and the screening conflict rate, leading to an overall decrease in 3.93 hours of weekly screening time. The improved efficiency did not come at the cost of accuracy, as precision and recall of the screening process and summary statistics of the included estimates were similar with and without the tool. Users provided positive feedback overall to using the tool.

Furthermore, one-person-one-tool and tool-only screening systems, which have the potential for increased time-reductions, performed as well as two human reviewers. The results of the OPOT analyses are particularly promising, as this system can reduce screening time by over 70%. Though the tool-only system performed comparably well to the OPOT system in our analyses, further study is required. Our pilot abstract sample was enriched with previously included texts, meaning that its precision at the abstract screening stage would likely be lower in practice. SeroTracker plans to adopt the OPOT(W) system into regular screening practice, to reduce screening time while maintaining a level of qualitycontrol through human monitoring. We will incorporate reviewer feedback to augment the tool’s utility further. The improvements above can ultimately improve the efficiency of the entire living systematic review, allowing us to maintain up-to-date information.

The failure cases of the tool provide avenues for future development. The most and least used features were the inclusion recommendation indicator and PIO highlighting, respectively. The performance of the named-entity-recognition used in PIO highlighting lags behind classification tasks in similar domains, [18] suggesting that more well-developed technologies may provide greater benefit. Furthermore, the tool did not aid with the full-text screening. As described in Section 3.1.2, 6 PI texts were excluded in full-text review in the with-tool stage. This suggests an inherent level of variation in the systematic review process that the tool could not mitigate.

In a broader context, our study supports previous reports on the potential for using NLP to improve screening efficiency without impacting review quality. We build off previous work by providing the first comprehensive analysis of the feasibility and impact of NLP implementation. We examined this technology within an ongoing living systematic review, rather than through a retrospective evaluation, and our study design allowed for documented user-specific aspects of NLP tool development. We addressed the research team’s specific needs when designing the tool’s features through our QI approach and evaluated how users interacted with them. User feedback demonstrated that NLP tools are useful to augment efficiency and that reviewers feel that they improved the screening workflow. Finally, the design of this project allowed us to compare characteristics of included texts in the systematic review with and without the use of this NLP tool, which has not been previously evaluated.

This project has implications for evidence synthesis at large and its role in evidencebased decision-making. As was highlighted by the COVID-19 pandemic, there are roadblocks to adequate evidence-based medicine during health emergencies. While the need for rapid research results is evident during a health emergency, evidence synthesis is generally slow and rigorous. [27] This incompatibility begets technological advancements, such as NLP-assisted tools, to accelerate the systematic review process. The success of this tool at SeroTracker demonstrates that these tools can fulfill this role and expedite the response of evidence-based medicine to a health emergency.

Furthermore, the living systematic review in particular provides an ideal context for the implementation of such tools, as results of an initial manual review can be used as a training set to develop a tool that would, in turn, expedite all future rounds of the review. Other teams working on living systematic reviews would benefit from integrating such tools tailored to their research question and design. Our QI project had a robust design, evaluated a comprehensive set of metrics, and can be used as a model. Appendix B provides more implementation details on the planning and technical aspects of the tool.

### 4.1 Limitations

Because the project was designed as a QI project in an ongoing review, the proportion of abstracts screened by individual reviewers varied from week to week. Reviewers may have systematic differences in how they vote, which could change the outcome of abstracts included. While the effect analysis of the tool on efficiency in Section 3.1.1 attempts to decompose the per-reviewer variability, there are other potentially informative covariates, such as whether the abstract received an “include” or “exclude” vote, or whether a reviewer had seen the abstract before, which were not accounted for.

Furthermore, the AB project design could induce order effects. While we showed in Section 3.1.1 that the removal of duplicate abstract-vote pairs did not affect the time decrease in abstract screening, we could not definitively demonstrate that order effects did not influence votes, particularly for texts that were accepted into full-text screening and subsequently interacted with multiple times. Finally, the sample size was limited to reduce screening load placed on the team, and some conclusions lacked the sample size for statistical significance. This screening load constraint also resulted in a higher inclusion rate in the pilot abstract set (23%) than what is typically observed in screening (5%).

Beyond design, there are limitations to assessing the impact of our tool as a whole. Firstly, there is no definitive “gold standard” for inclusion or exclusion of abstracts; we assumed “pre-project” screening labels to be accurate. Furthermore, our precision and recall analyses treated all false negatives as equal value, but studies generally contribute differently to the quantitative results of a review. For instance, falsely excluding a paper reporting several unique seroprevalence estimates would alter overall results more than excluding a paper with just one estimate. While we examined summary statistics of missed texts in Section 3.2.1, a meta-analysis was not performed to determine the impact on downstream analysis. Finally, since the tool incorporated several features, the individual contributions of each feature could not be quantified. While we assessed user interaction and agreement with individual features in Section 3.2.2, the number of covariates, due to the multiple features, prevent this analysis from demonstrating causality.

## 5 Conclusion

This study provides the first comprehensive analysis on the implementation of NLP technology in the screening stage of a living systematic review. Incorporating an internally developed tool at SeroTracker was feasible and significantly improved our processes by increasing screening efficiency while maintaining accuracy. User feedback was positive, leading to continued use of the tool in regular practice. This provides promising data for the evidence synthesis community, as similar tools could be used to expedite the timeconsuming and laborious manual systematic review process for other research groups, allowing for more up-to-date dissemination of evidence syntheses.

## Data Availability

All data produced in the present work are contained in the manuscript text or at the links provided in the manuscript

https://github.com/yolky/Serotracker-NLP-Tool-Analysis

https://github.com/serotracker/Serotracker-NLP-Training-and-Inference

## Acknowledgements

We greatly thank the reviewers on the SeroTracker team, including Mercedes Yanes-Lane, Anabel Selemon, Natasha Ilincic, Judy Chen, Christian Cao, Mairead Whelan, Zihan Li and Xiaomeng Ma for their participation in the quality improvement project. We also thank Gabriel Deveaux, Himanshu Ranka, and Aaron Bensmihen for help with a review of the finalized manuscript.

## Funding Information

SeroTracker receives funding for SARS-CoV-2 seroprevalence study evidence synthesis from the Public Health Agency of Canada through Canada’s COVID-19 Immunity Task Force, the World Health Organization Health Emergencies Programme, the Robert Koch Institute, and the Canadian Medical Association Joule Innovation Fund. No funding source had any role in the design of this study, its execution, analyses, interpretation of the data, or decision to submit results. This manuscript does not necessarily reflect the views of the World Health Organization or any other funder.

## Conflicts of Interest

RKA was previously a Technical Consultant for the Bill and Melinda Gates Foundation Strategic Investment Fund, is a minority shareholder of Alethea Medical, and was a former Senior Policy Advisor at Health Canada. Each of these relationships is unrelated to the present work. TY reports a role at the Centre for Addiction and Mental Health and past employment at Health Canada, outside of the submitted work. All other authors declare that they have no competing interests.

## Author Contributions

NL contributed to the development of the NLP tool. SPA, NL, and RKA contributed to the investigation and data collection. NL contributed to the statistical analysis and visualization. SPA and NL contributed to writing of the manuscript. RKA, NB, and TY contributed to the conceptualisation, methodology, and review and editing of the manuscript. All authors approved of the final manuscript prior to submission.

## Data Availability

Code for data analysis performed in this study as well as code for the training of the NLP tool are published in public Github repositories [28, 29].

## Footnotes

1 If, during full-text extraction, a text that was included in screening is found to lack information to extract, it can be excluded

## References

1. Garritty C, Stevens A, Hamel C, Golfam M, Hutton B, Wolfe D. Knowledge Synthesis in Evidence-Based Medicine. Semin Nuclear Med.. 2019;49(2):136–144. doi: 10.1053/j.semnuclmed.2018.11.006.

2. Guyatt GH, Sackett DL, Sinclair JC, et al. Users’ Guides to the Medical Literature: IX. A Method for Grading Health Care Recommendations. JAMA.. 1995;274(22):1800– 1804. doi: 10.1001/jama.1995.03530220066035.

3. COVID-19 Primer. https://covid19primer.com/. Accessed October 6, 2021.

4. Borah R, Brown AW, Capers PL, Kaiser KA. Analysis of the Time and Workers Needed to Conduct Systematic Reviews of Medical Interventions Using Data from the PROSPERO Registry. BMJ Open.. 2017;7(2):e012545. doi: 10.1136/bmjopen-2016-012545.

5. Bastian H, Glasziou P, Chalmers I. Seventy-Five Trials and Eleven Systematic Reviews a Day: How Will We Ever Keep Up?. PLoS Med.. 2010;7(9):e1000326. doi: 10.1371/journal.pmed.1000326.

6. Pearson H. How COVID Broke the Evidence Pipeline. Nature.. 2021;593(7858):182– 185. doi: 10.1038/d41586-021-01246-x.

7. Elliott JH, Turner T, Clavisi O, et al. Living Systematic Reviews: An Emerging Opportunity to Narrow the Evidence-Practice Gap. PLoS Med.. 2014;11(2):e1001603. doi: 10.1371/journal.pmed.1001603.

8. Millard T, Synnot A, Elliott J, Green S, McDonald S, Turner T. Feasibility and Acceptability of Living Systematic Reviews: Results from a Mixed-Methods Evaluation. Syst Rev.. 2019;8(1):325. doi: 10.1186/s13643-019-1248-5.

9. Marshall IJ, Wallace BC. Toward Systematic Review Automation: A Practical Guide to Using Machine Learning Tools in Research Synthesis. Syst Rev.. 2019;8(1):163. doi: 10.1186/s13643-019-1074-9.

10. Chai KEK, Lines RLJ, Gucciardi DF, Ng L. Research Screener: A Machine Learning Tool to Semi-Automate Abstract Screening for Systematic Reviews. Syst Rev.. 2017;10(1):93. doi: 10.1186/s13643-021-01635-3.

11. Gates A, Guitard S, Pillay J, et al. Performance and Usability of Machine Learning for Screening in Systematic Reviews: A Comparative Evaluation of Three Tools. Syst Rev.. 2019;8(1):278. doi: 10.1186/s13643-019-1222-2.

12. Hamel C, Kelly SE, Thavorn K, Rice DB, Wells GA, Hutton B. An Evaluation of DistillerSR’s Machine Learning-Based Prioritization Tool for Title/Abstract Screening – Impact on Reviewer-Relevant Outcomes. BMC Med Res Methodol.. 2020;20(1):256. doi: 10.1186/s12874-020-01129-1.

13. Lerner I, Créquit P, Ravaud P, Atal I. Automatic Screening Using Word Embeddings Achieved High Sensitivity and Workload Reduction for Updating Living Network Meta-Analyses. J Clin Epidemiol.. 2019;108:86–94. doi: 10.1016/j.jclinepi.2018.12.001.

14. Ouzzani M, Hammady H, Fedorowicz Z, Elmagarmid A. Rayyan—a Web and Mobile App for Systematic Reviews. Syst Rev.. 2016;5(1):210. doi: 10.1186/s13643-016-0384-4.

15. Wallace BC, Trikalinos TA, Lau J, Brodley C, Schmid CH. Semi-Automated Screening of Biomedical Citations for Systematic Reviews. BMC Bioinformatics.. 2010;11(1):55. doi: 10.1186/1471-2105-11-55.

16. Qin X, Liu J, Wang Y, et al. Natural Language Processing Was Effective in Assisting Rapid Title and Abstract Screening When Updating Systematic Reviews. J Clin Epidemiol.. 2021;133:121–129. doi: 10.1016/j.jclinepi.2021.01.010.

17. Devlin J, Chang MW, Lee K, Toutanova K. BERT: Pre-Training of Deep Bidirectional Transformers for Language Understanding. CoRR. 2018;abs/1810.04805 http://arxiv.org/abs/1810.04805.

18. Gu Y, Tinn R, Cheng H, et al. Domain-Specific Language Model Pretraining for Biomedical Natural Language Processing. ACM Trans. Comput. Healthcare.. 2022;3(1):1–23. doi: 10.1145/3458754.

19. Huang KC, Chiang IJ, Xiao F, Liao CC, Liu CCH, Wong JM. PICO Element Detection in Medical Text without Metadata: Are First Sentences Enough?. J. Biomed. Inform.. 2013;46(5):940–946. doi: 10.1016/j.jbi.2013.07.009.

20. Cohen AM, Hersh WR, Peterson K, Yen PY. Reducing Workload in Systematic Review Preparation Using Automated Citation Classification. J Am Med Inform Assoc.. 2006;13(2):206–219. doi: 10.1197/jamia.M1929.

21. Schoot R, Bruin J, Schram R, et al. An Open Source Machine Learning Framework for Efficient and Transparent Systematic Reviews. Nat Mach Intell.. 2021;3(2):125–133. doi: 10.1038/s42256-020-00287-7.

22. Arora RK, Joseph A, Van Wyk J, et al. SeroTracker: A Global SARS-CoV-2 Seroprevalence Dashboard. Lancet Infect Dis.. 2021;21(4):e75–e76. doi: 10.1016/S1473-3099(20)30631-9.

23. Bobrovitz N, Arora RK, Cao C, et al. Global Seroprevalence of SARS-CoV-2 Antibodies: A Systematic Review and Meta-Analysis. PLOS ONE.. 2021;16(6):e0252617. doi: 10.1371/journal.pone.0252617.

24. Ltd VHI. Covidence. https://www.covidence.org/. Accessed November 5, 2021.

25. Nye B, Li JJ, Patel R, et al. A Corpus with Multi-Level Annotations of Patients, Interventions and Outcomes to Support Language Processing for Medical Literature. in Proceedings of the 56th Annual Meeting of the Association for Computational Linguistics (Volume 1: Long Papers):197–207.Association for Computational Linguistics. 2018.

26. Wang Z, Nayfeh T, Tetzlaff J, O’Blenis P, Murad MH. Error Rates of Human Reviewers during Abstract Screening in Systematic Reviews. PLOS ONE.. 2020;15(1):e0227742. doi: 10.1371/journal.pone.0227742.

27. Evidence-Based Medicine: How COVID Can Drive Positive Change. Nature.. 2021;593(7858):168–168. doi: 10.1038/d41586-021-01255-w.

28. SeroTracker. Serotracker-NLP-Tool-Analysis. Github. https://github.com/serotracker/Serotracker-NLP-Tool-Analysis 2021. Accessed December 24, 2021.

29. SeroTracker. Serotracker-NLP-Training-and-Inference. Github. https://github.com/serotracker/Serotracker-NLP-Training-and-Inference 2022. Accessed January 4, 2022.

