## Appendix for "A real-world evaluation of the implementation of NLP technology in abstract screening of a systematic review"

### A Additional Results

#### A.1 Voter Agreement with Inclusion Recommendations

We assessed voter agreement with the tool's recommendations. Figure A1b shows how the precision and recall varies as the recommendation threshold varies, while Figure A1a shows the resulting ROC curve, where the "true label" is given by the user's vote, and the comparison is whether the tool's recommendation is at least above the given recommendation threshold. These votes include all abstracts in the with-tool stage. The ROC curve in Figure A1a yields an AUC of 0.956, suggesting that users very strongly voted in alignment with the tools recommendations, most in alignment with the "somewhat recommended" threshold.

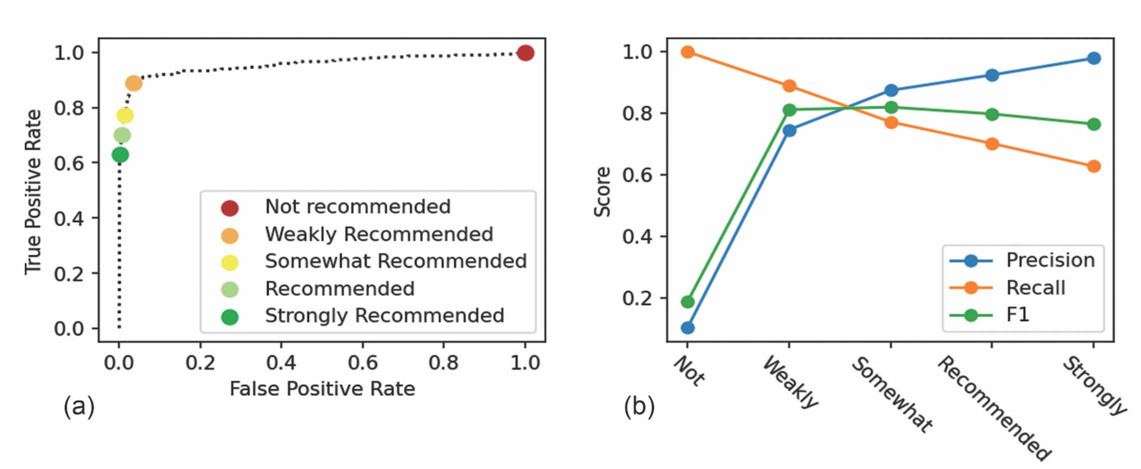


**Figure A1:** Operating characteristics of the tool comparing recommendations given by the tool at the five confidence levels to "true labels" given by user votes. (a) shows the operating curve and five confidence levels, while (b) shows how the precision, recall and F1 score varies with the change confidence.

#### A.2 Voter Alignment with the Presence of Keywords

We found a positive correlation between keywords and user's votes: 83.8% (109/130) of the include votes contained keywords in the abstracts, compared with only 37.7% (429/1137) of the exclude votes (p=2.29e-24, Fisher's exact test). For texts which did not meet the lowest recommendation threshold, 71.4% (10/14) of texts that received include votes had keywords compared to 37.1% (412/1112) that received exclude votes (p=0.011, Fisher's exact test). This suggests that keywords could be useful signal to reviewers in cases where the inclusion recommendation feature does not recommend a text. The most common keywords were terms associated with inclusion, such as "seroprevalence," "antibody," and "serology," suggesting that users were primarily using keywords as a positive indicator rather a negative one.

### B Training and Development of the Tool

#### B.1 Dataset partitioning

Our entire dataset of classified texts consisted of 38674 abstracts, as of early July 2021, when the model was trained. Since we were interested in classification based on the abstracts, we ignored all texts which did not contain an abstract. Of these remaining abstracts, 3.4% of them were moved to the extraction stage (included), 1.4% were rejected in full text review and the remaining 95.2% were deemed ineligible in abstract screening. We split our data into three categories: the pilot abstracts, the validation set, and the training set.

For selecting pilot abstracts, we looked at the subset of abstracts that were screened in April 2021. This period was chosen so that it was close enough to the project time frame for texts to not be obviously outdated relative to the newly uploaded ones, but not so recent that reviewers could remember them. From this set, 100 abstracts which were included in full text review were randomly sampled. Then, for the remaining 300 abstracts, we uniformly sampled from the combined set of abstracts which were rejected in abstract and full text screening. Of note, this represents a substantially increased positive-negative ratio compared to a simple random sample of texts; in particular the number of texts rejected in full-text screening is smaller than would be expected relative to the number of accepted texts. The positivity rate was purposefully increased, in order to produce a large enough sample of included texts, without requiring a large overall pilot set for the team to re-screen. Since the pilot abstracts were mixed with new abstracts during screening, the positivity rate of the overall set was not drastically changed. After further inspection of the 100 abstracts that were included, we found that 9 of them were actually excluded at the extraction stage. Thus, we classified them as negatives, resulting in 91 included texts and 309 excluded texts in our pilot abstracts.

Of the remaining 33783 abstracts, 30% of them were set aside as a validation set, on which no models were trained with which model selection was done. The remaining 70% was used in the training set.

#### B.2 Data pre-processing

We concatenated the title followed by the abstract together to be fed into the model. We removed commonly found html tags and escape characters found in the abstracts such as “<h4>” or “\t.” If the resulting string exceeded 512 tokens (the maximum for the BERT model), we simply removed any excess. Because we had a highly imbalanced dataset, we found that upsampling the positive samples in the training set by a factor of 25 significantly improved recall rate, at the cost of some precision. Due to the relative cost of missing abstracts in text screening, we found this trade-off worthwhile. We found that other common NLP text augmentation techniques such as random sentence cropping, text flipping, or synonym swapping produced minor performance gains which were not robust to changes in other training hyperparameters, so we did not use these.

#### B.3 Training

We trained an ensemble of 5 models on random samples of 60% of the training set. Each model was a fine-tuned version of PubMedBERT (uncased, abstracts only). We trained for 4 epochs with batch sizes of 32 and a learning rate of 1e-5. The predictions of the five models were averaged together to form the final inclusion likelihood.

All code used to train the tool can be found at the following link: https://github.com/serotracker/Serotracker-NLP-Training-and-Inference

#### B.4 Training Results

Table B1 summarizes the performance of the tool evaluated on the validation set for the various confidence thresholds. We additionally report the 50% threshold, which is the default for most classification tasks. At this threshold, the F1 score was .818, with a better recall of .871. At the lowest recommendation threshold, we achieved a recall of .963, at a precision of .406, meaning that nearly all abstracts are captured at this lowest threshold. Figure B2 shows the resulting ROC and calibration curve. We also visualize the four recommendation thresholds, corresponding to 15%, 35%, 60%, and 75% predictions by the classifier. As seen by the calibration curve, the inclusion likelihoods are not well calibrated, which motivated the use of these thresholds as opposed to directly reporting the score to the user. We achieve an AUC of 0.987, which is slightly better than 0.956 AUC found when comparing the tool to user's votes. This is expected, as the model was chosen to perform well on this validation set.

**Table B1:** Operating characteristics of the tool at different confidence thresholds on the test set of abstracts.

| | TN | FP | FN | TP | Prec. | Rec. | F1 |
|---|---|---|---|---|---|---|---|
| Weakly Recommended | 9416 | 416 | 11 | 284 | 0.406 | 0.963 | 0.571 |
| Somewhat Recommended | 9718 | 114 | 29 | 266 | 0.700 | 0.902 | 0.788 |
| Recommended | 9775 | 57 | 41 | 254 | 0.817 | 0.861 | 0.838 |
| Strongly Recommended | 9801 | 31 | 61 | 234 | 0.883 | 0.793 | 0.836 |
| 50% Threshold (For Development) | 9756 | 76 | 38 | 257 | 0.772 | 0.871 | 0.818 |

Abbreviations: TN, true negative ; FP, false positive ; FN, false negative ; TP, true positive ; Prec., precision ; Rec., recall

##### B.5 PIO Highlighting

For PIO higlighting, we trained the model using the EBM-NLP dataset for PICO extraction. Again, we used the same PubMedBERT model as the base. Because abstracts were often longer than 512 tokens, for PIO higlighting we split the abstracts into sets of sentences at most 512 tokens long, and fed these sets into the model separately, and combined the results of the two sets. We found that there was a tendency to often repeatedly highlight blocks of text which were semantically similar. Thus, we trained a separate model on the MedSTS dataset, a dataset for sentence similarity. We used a cosine similarity threshold of 0.95 for removing PIO highlights which were deemed too similar by that model. Finally, we found that the model would produce far too many highlights, often of very short strings. In the plugin we only highlight the top three highlighted blocks for each category in population, intervention and outcome, measured by the probability assigned to the highlight, averaged over a contiguous highlighted block.

#### C Infrastructure and Implementation

This section covers implementation details related to the infrastructure and setup of the tool from a practical standpoint, for the benefit of other teams looking to implement something

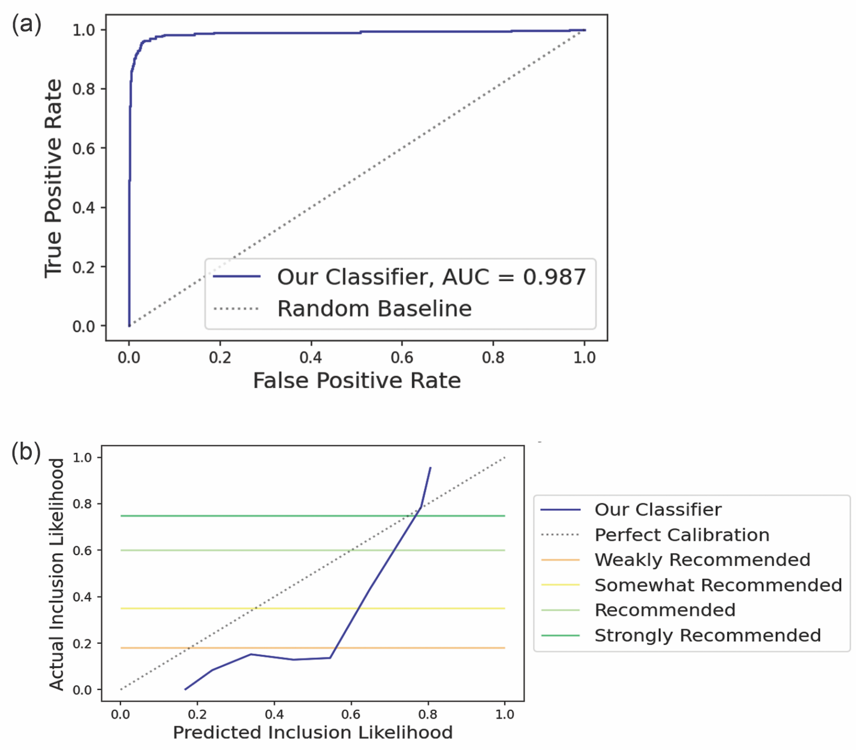


**Figure B2:** (a) ROC and (b) calibration curve of the inclusion recommendation system evaluated on the set of validation abstracts.

similar. There are two main software components of the tool: the plugin itself, and a server which produces the predictions. We briefly outline the design and some practical considerations for those who look to produce something similar.

#### C.1 The Chrome Plugin

This plugin was developed in javascript as a add-on for chrome. Each member of the team had to manually install the plugin. In Figure C3 we show how the plugin interacts with the Covidence page. Each feature can be individually turned on or off. All of the NLP-based features (namely inclusion recommendations and PIO highlighting) are served a separate server, as opposed to generated locally.

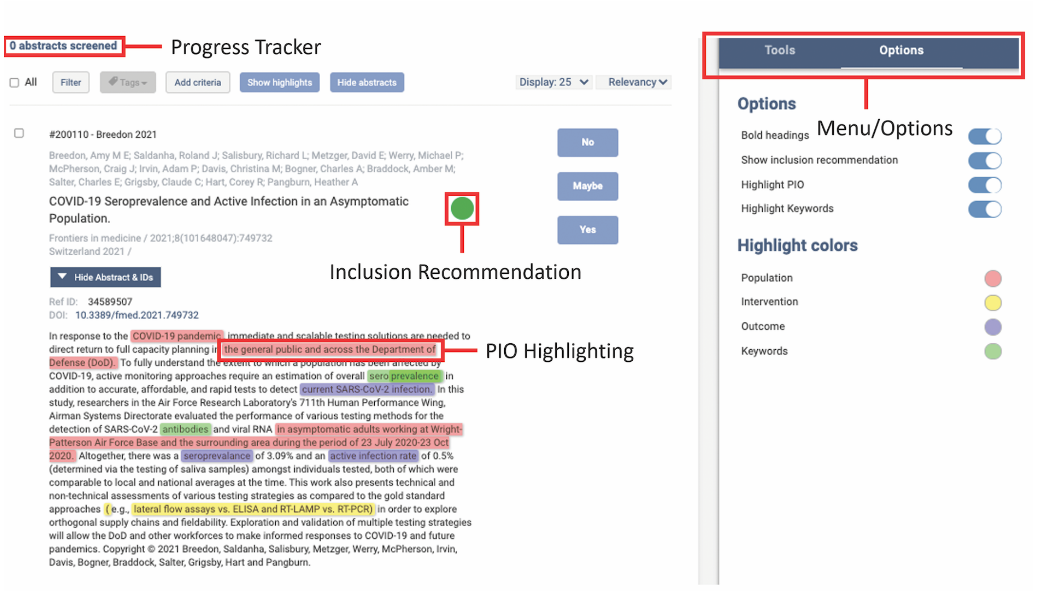


**Figure C3:** An example of how the plugin interacts with an abstract on Covidence, including the key features: progress tracking, inclusion recommendations, and PIO highlighting are shown.

#### C.2 The Server

To generate the predictions, we used a separate AWS server. Because inference requires substantial compute power, rather than that generating predictions in real-time, we simply used the server as a database to store the pre-generated results. Particularly, after each weekly upload of new abstracts, we generate inclusion recommendations and PIO highlights for all of the abstracts. This process generally takes around 8 minutes on a machine with an Nvidia RTX 3090. These predictions are sent to the server and stored in the database. When users interact with the chrome plugin, a request is sent to the server to read from the predictions database. This choice to cache results reduces the latency for predictions from around 30 seconds to several milliseconds, compared to generating predictions in real time. We strongly advise this approach if it is possible.

### D Survey on Usefulness of the Tool's Features

Following the project, a questionnaire was sent to team members in the form of a google form, asking for feedback on the tool and its features. Feedback was provided anonymously. The contents of the questionnaire are outlined below:

**SECTION 1:USEFULNESS OF THE TOOL'S FEATURES**: Please answer the following questions about how useful you found the tool's features. By "useful" we mean how helpful you found these features at improving screening, by making the process faster and/or easier for you in any way.

1) How useful did you find the progress tracking feature on a scale from 1 to 5, where 1 is "not useful at all" and 5 is "extremely useful"? By "useful", we mean how helpful you found this feature at improving screening, by making the process faster and/or easier for you in any way.

2) Which of the optional features of the NLP plugin did you use while screening? (Select all that apply)

- The undo feature
- The inclusion recommendations feature
- The automatic bolding of headings in the abstract (i.e. the bolding of headings such as "Background" and "Methods"
- The keyword highlight feature
- The PIO (Population, Intervention, Outcome) highlight feature

3) If you used the undo feature, how useful did you find it on a scale from 1 to 5, where 1 is “not useful at all” and 5 is “extremely useful”? (If you did not use this feature, please leave blank)

4) If you used the inclusion recommendations feature, how useful did you find it on a scale from 1 to 5, where 1 is “not useful at all” and 5 is “extremely useful”? (If you did not use this feature, please leave blank)

5) If you used the bolding of headings feature, how useful did you find it on a scale from 1 to 5, where 1 is “not useful at all” and 5 is “extremely useful”? (If you did not use this feature, please leave blank)

6) If you used the keyword highlight feature, how useful did you find it on a scale from 1 to 5, where 1 is “not useful at all” and 5 is “extremely useful”? (If you did not use this feature, please leave blank)

7) If you used the keyword highlight feature, which keywords would you typically put in the box?

8) If you used the PIO highlight feature, how useful did you find it on a scale from 1 to 5, where 1 is “not useful at all” and 5 is “extremely useful”? (If you did not use this feature, please leave blank)

**SECTION 2: OVERALL EXPERIENCE**: Please answer the following questions about your overall experience with the tool. When answering, please keep in mind the usefulness of the tool in itself, and not the design of the project we conducted.

1) Overall, did using this tool during screening make the screening process faster and more efficient for you?

- Yes
- No

2) Overall, did using this tool during screening make the screening process easier for you?

- Yes
- No

3) Overall, did using this tool during screening improve the screening process?

- Yes
- No

4) Please provide additional comments on why/how you felt the tool was beneficial. Some questions to consider are: Which features did you find most useful and why? Which aspects of the overall screening process did the tool improve for you? Specifically, in which way was the process improved (eg. efficiency, ease of screening, etc.) and why? If you have any additional comments on the positive aspects of the tool, you may add them here.

5) Please provide additional comments on why/how you felt the tool was NOT beneficial. Some questions to consider are: Are there any features that were not useful at all? If yes, which ones and why? Did the tool detract from the quality of the screening process

in any way? If yes, please elaborate on why this was the case. If you have any additional comments on the negative aspects of the tool, you may add them here.

6) Building off your responses to the previous question, please elaborate on how the tool could be improved. Some questions to consider are: If there were any features of the tool you felt were not useful, how could these features be improved? Are there any additional features that you feel would further benefit the tool? If the tool detracted from the screening process in any way, how can this be avoided?

7) Would you like to continue using this tool during screening?

- Yes
- No
- Only if certain changes are made

8) If you checked off “only if certain changes are made” above, please explain what these changes are.
